# Depressive symptoms and incident osteoporosis: Findings from a 20-year follow up of the English Longitudinal Study of Ageing

**DOI:** 10.64898/2026.07.27.26359005

**Authors:** Ruth A. Hackett, James B. Galloway, Mark D. Russell, Lydia Poole, Amy Ronaldson, Sam Norton

**Affiliations:** Health Psychology Section, Department of Psychology, Institute of Psychiatry, Psychology and Neuroscience, King’s College London, London, UK; Centre for Rheumatic Disease, Department of Inflammation Biology, School of Immunology & Microbial Sciences, King’s College London, London, UK; Centre for Psychiatry and Mental Health, Wolfson Institute of Population Health, Queen Mary University of London, London, UK; Department of Health Service and Population Research, Institute of Psychiatry, Psychology and Neuroscience, King’s College London, London, UK

**Keywords:** Depression, osteoporosis, mid-age, older age, cohort study

## Abstract

**Purpose:** Psychosocial factors are underexplored in relation to osteoporosis. This study examined whether depressive symptoms are associated with incident osteoporosis in middle aged and older adults, and whether this association is modified by age or sex.

**Methods:** Prospective cohort study of 8,577 adults aged ≥50 years from the English Longitudinal Study of Ageing. The sample were free of osteoporosis at baseline (2002–2003) and were followed up for incident osteoporosis until wave 11 (2023–2024). Depressive symptoms were measured at baseline (2002-03) using the 8-item Center for Epidemiologic Studies-Depression Scale (CES-D). Cox proportional hazards regressions were used to estimate associations, adjusting for age, sex and wealth.

**Results:** Over 20 years of follow-up, 948 (11.1%) participants reported incident osteoporosis. Each one-point increase in baseline CES-D was associated with a higher hazard of incident osteoporosis adjusted for age, sex, and wealth (hazard ratio (HR) 1.07, 95% CI 1.04–1.11, *p*<0.001). The association was modified by age (interaction *p*=0.008) and concentrated in participants aged 50–64 years at baseline (HR 1.10, 95% CI 1.06–1.14), with no significant association in older age strata. Osteoporosis incidence was higher in women, but this was not modified by depressive symptoms. Findings were robust to sequential adjustment (informed by a directed acyclic graph) for education, wealth, inflammatory conditions and health behaviours (HR 1.04, 95% CI 1.01-1.08)

**Conclusion:** Depressive symptoms are associated with incident osteoporosis over two decades of follow-up, particularly among adults aged 50–64. Mid-life may represent a target for psychosocial intervention to support bone health.

**Mini abstract:** Depressive symptoms are a plausible psychosocial risk factor for osteoporosis, but longitudinal evidence is scarce. In 8,577 adults followed for 20 years, higher depressive symptoms were associated with incident osteoporosis, concentrated in those aged 50 to 64. Mid-life depressive symptoms may be a modifiable target for protecting later bone health.

## 1. Introduction

Osteoporosis is a chronic condition characterised by weakened bones and increased fracture risk, representing a major public health challenge as ageing populations expand globally. In the United Kingdom, approximately 3.77 million adults (5.2% of the population) had osteoporosis in 2019, with women representing 78.3% of cases[1]. Osteoporosis develops slowly, is largely asymptomatic, and is often only detected following a fracture. More than 500,000 fragility fractures occur annually in the UK, causing pain, disability and reduced quality of life, while accounting for approximately 2.4% (£5.4 billion) of NHS spending each year[1, 2].

Despite this burden, osteoporosis remains under-researched relative to other chronic conditions[3]. Psychosocial factors such as depressive symptoms are well-established risk factors for other long-term conditions[4] including cardiovascular disease[5], type-2 diabetes[6] and rheumatoid arthritis[7], yet remain comparatively underexplored in the context of osteoporosis.

Several plausible mechanisms could link depressive symptoms to osteoporosis[8]. Depressive symptoms are associated with adverse health behaviours including physical inactivity (which reduces mechanical loading on bone)[8, 9], smoking (direct toxic effects on bone metabolism)[8, 10] and excess alcohol consumption (suppressing osteoblast activity)[11]. Direct biological pathways including chronic inflammation may also contribute[12, 13]. However, the existing literature linking depressive symptoms with osteoporosis is sparse.

Earlier work relies predominantly on small nonrepresentative samples with a focus on bone mineral density rather than osteoporosis diagnosis[8]. Recent studies in larger representative samples have been mostly cross-sectional[14–18] or have focused on other outcomes such as fracture[19, 20] rather than incident osteoporosis. For example, a 2026 cross-sectional analysis examined frailty-depression-osteoporosis co-occurrence in England and the US[14]. Three recent US-based studies on co-occurring depression and osteoporosis[15–17], and one on history of psychological disorders and osteoporosis[18] all reported significant associations but note the limited evidence base and need for longitudinal studies.

Overall, there is a paucity of large prospective cohort studies examining depressive symptoms as a predictor of incident osteoporosis in representative ageing populations. Furthermore, given that both osteoporosis risk[1] and depressive symptom prevalence[21] vary by age and sex, it is important to examine whether associations are uniform across demographic groups or are concentrated within specific subgroups.

The current study aimed to address these gaps using data from the English Longitudinal Study of Ageing (ELSA), a representative panel study of adults aged 50 years and older. Our primary objective was to determine whether depressive symptoms at baseline associate with greater risk of incident osteoporosis over 20 years of follow-up. Secondary objectives were to examine whether the association is modified by age or sex, and to explore the extent to which behavioural pathways (physical activity, alcohol consumption, smoking status) account for any observed association.

## 2. Methods

### 2.1 Participants

ELSA is a representative panel study of community-dwelling adults aged 50 years and older living in England. Data collection began in 2002–03 (wave 1), with follow-up waves at two-yearly intervals[22]. Each wave, data are collected via computer-assisted personal interview and self-reported questionnaires. ELSA received ethical approval from the London Multicentre Research and Ethics Committee (MREC/01/02/91) in accordance with the World Medical Association Declaration of Helsinki. All participants provided informed consent.

### 2.2. Predictor variable: Depressive symptoms

Depressive symptoms were measured using the 8-item Center for Epidemiologic Studies-Depression Scale (CES-D)[23]. Participants gave dichotomous (yes/no) responses to items such as *’I felt depressed’* and *’My sleep was restless’,* producing a total score ranging from 0 to 8, with higher scores indicating greater depressive symptoms. The CES-D was modelled as a continuous score for the main analysis, while a cut-point of ≥3 indicating raised depressive symptoms[4] was used for graphical purposes (Kaplan-Meier curve).

### 2.3 Outcome variable: Incident osteoporosis

Each wave, participants were asked whether a physician had diagnosed them with osteoporosis since their last interview. Time of diagnosis was indexed as the wave at which osteoporosis was first reported. Time to event was measured in years from wave 1 (2002-03). For participants not diagnosed with osteoporosis by their last observed wave, time was censored at the last wave of participation, defined using wave-specific age variables (waves 2-11) as an indicator of survey participation.

### 2.4 Covariates

Covariates were measured at baseline (wave 1, 2002-03). Participants self-reported their age (years), sex (male/female), and non-pension household wealth (divided into quintiles across the wave 1 sample). Wealth was selected as the primary indicator of socio-economic position, as it has been identified as the most relevant marker for this cohort, as it most sensitivity reflects the material circumstances of middle aged and older adults[22]. Age, sex and wealth were included as the primary covariate set across models.

In additional analyses informed by a directed acyclic graph (DAG; Supplementary Figure 1), we considered additional covariates measured at baseline. We explored education as an alternative indicator of socio-economic position, as wealth was measured concurrently with depressive symptoms it may lie on a pathway shared with osteoporosis, whereas education is more reflective of past socio-economic position so is not subject to this concern. Education was self-reported and derived as a 4-level variable (0=no qualification, 1= foreign/other qualification. 2=qualification below university degree; 3= university degree or higher). We also considered other chronic illnesses with an inflammatory component which could confound the relationship between depressive symptoms and osteoporosis: self-reported chronic lung disease (yes/no), arthritis (yes/no). and asthma (yes/no). Finally, we explored behavioural pathways, by further adjustment for three self-reported behavioural variables: physical activity (5-category scale ranging from sedentary to active), alcohol use (none vs. any), and smoking status (never/former/current). We hypothesised body mass index (BMI) and history or trait depressive symptoms were on the causal pathway from depressive symptoms to osteoporosis [24]. However, as these were unmeasured at the wave 1 baseline, we were unable to include these in our models.

### 2.5 Statistical analysis

Descriptive characteristics are presented as mean (standard deviation) or number (percentage). The characteristics of those who did and did not develop incident osteoporosis were compared using *t-*tests and chi-squared tests. Cox proportional hazards regression was used to investigate the association between baseline depressive symptoms and incident osteoporosis. The proportional hazards assumption was assessed by including a time-by-exposure interaction term (CES-D × log time) and was not violated (*p* = 0.155). Hazard ratios (HR) and 95% confidence intervals (CIs) represent the change in hazard per one-point increase in CES-D score. Our models adjusted for age, sex and non-pension wealth quintile. The CES-D was modelled as a continuous score for the main analysis, while a cut-point of 3 indicating raised depressive symptoms was used for the Kaplan-Meier curve. As a sensitivity check, models were re-run excluding participants with incident osteoporosis recorded at wave 2 (i.e. within two years of depressive symptoms assessment, *n* = 189) to address potential reverse causation. That is, for potential bias due to osteoporosis being present but not yet diagnosed at the time depressive symptoms were assessed. We did not have access to linked ELSA mortality data, so it was not possible to account for death as a competing risk. As a sensitivity analysis, we used multiple imputation to retain participants excluded from the primary analysis due to missing covariate data. The imputation sample (*n* = 9,074) comprised the complete-case sample (*n* = 8,577) plus 497 participants (5.5%) who were ≥50 years, osteoporosis-free at baseline and provided follow-up data but had missing CES-D, sex or wealth data. Participants without follow-up were ineligible for imputation. Missing covariates were imputed by chained equations (20 imputations), with the event indicator, Nelson-Aalen cumulative hazard and covariates included to remain compatible with the Cox model[25]. Estimates were pooled using Rubin’s rules.

To assess effect modification by age and sex we added multiplicative interaction terms to our models (CES-D × age; CES-D × sex). Where an interaction was significant, stratified models were planned by pre-specified age bands (50–64, 65–74, 75+ years) for age interactions and by sex (male/female) for sex interactions. In DAG informed analyses (Supplementary Figure 1), we assessed possible attenuation by covariates on the hypothesised causal pathway from depressive symptoms to osteoporosis among the 8,212 participants (926 incident cases) with complete data on education, chronic lung disease, arthritis, asthma, physical activity, alcohol use, and smoking status. Within this sub-sample, we compared the CES-D coefficient from a model adjusted for age, sex and education (Model 1) with the same model additionally adjusted for wealth and the three inflammatory conditions (Model 2), and finally a model adjusted for the aforementioned sociodemographic and inflammatory condition variables with the three behavioural covariates (Model 3). Percentage attenuation was calculated on the log hazard scale ((B_reference − B_behaviour) / B_reference × 100). Analyses were conducted in SPSS version 31. Claude/Anthropic was used to troubleshoot analytic code and help prepare tables.

## 3. Results

### 3.1 Participant characteristics

In this study we investigated the association between depressive symptoms measured at wave 1 (2002–03) and incident osteoporosis from wave 2 (2004–05) to wave 11 (2023–24), providing up to 20 years of follow-up. A flowchart of those included and excluded in the analysis is provided in Supplementary Figure 2. Analyses were restricted to core ELSA participants aged 50 years or older at baseline (*n =* 11,522), consistent with previous ELSA analyses of depressive symptoms and incident disease[4]. Participants were included if they reported no doctor-diagnosed osteoporosis at wave 1 (*n* = 10,957), had complete wave 1 data on depressive symptoms (*n* = 10,531) and age, sex and wealth (*n* = 10,250). A total of, 8,577 participants provided one wave of follow-up data, and these participants constituted the final analytic sample.

Participant characteristics are presented in Table 1. Compared with those excluded from the analyses (*n* = 3,522), participants included in the study (*n* = 8,577) were less likely to be female (53.2% vs 62.4%), wealthier (lowest wealth quintile: 17.2% vs 26.7%), more educated (degree: 12.1% vs 10%) and reported fewer depressive symptoms (mean 1.49 vs 1.83) at baseline (all *p* < 0.001). They were also more physically active (sedentary: 15.6% vs 31.8%, *p* < 0.001), more likely to consume alcohol (71.1% vs 63.5%, *p* < 0.001), and less likely to be current smokers (17.4% vs 19.3%, *p* = 0.036). The included group were significantly less likely to report asthma (11% vs 13.4%, *p* < 0.001) or lung disease (5.6% or 7.9%, *p* < 0.001) These differences are typical of complete case analysis of ELSA and other cohort studies[26]. Fewer female participants in the included sample likely reflect the exclusion of those with prevalent osteoporosis at baseline (*n*=565) given the higher prevalence of disease in women. The excluded and included group did not differ in arthritis prevalence (*p* = 0.943)

**Table 1:**
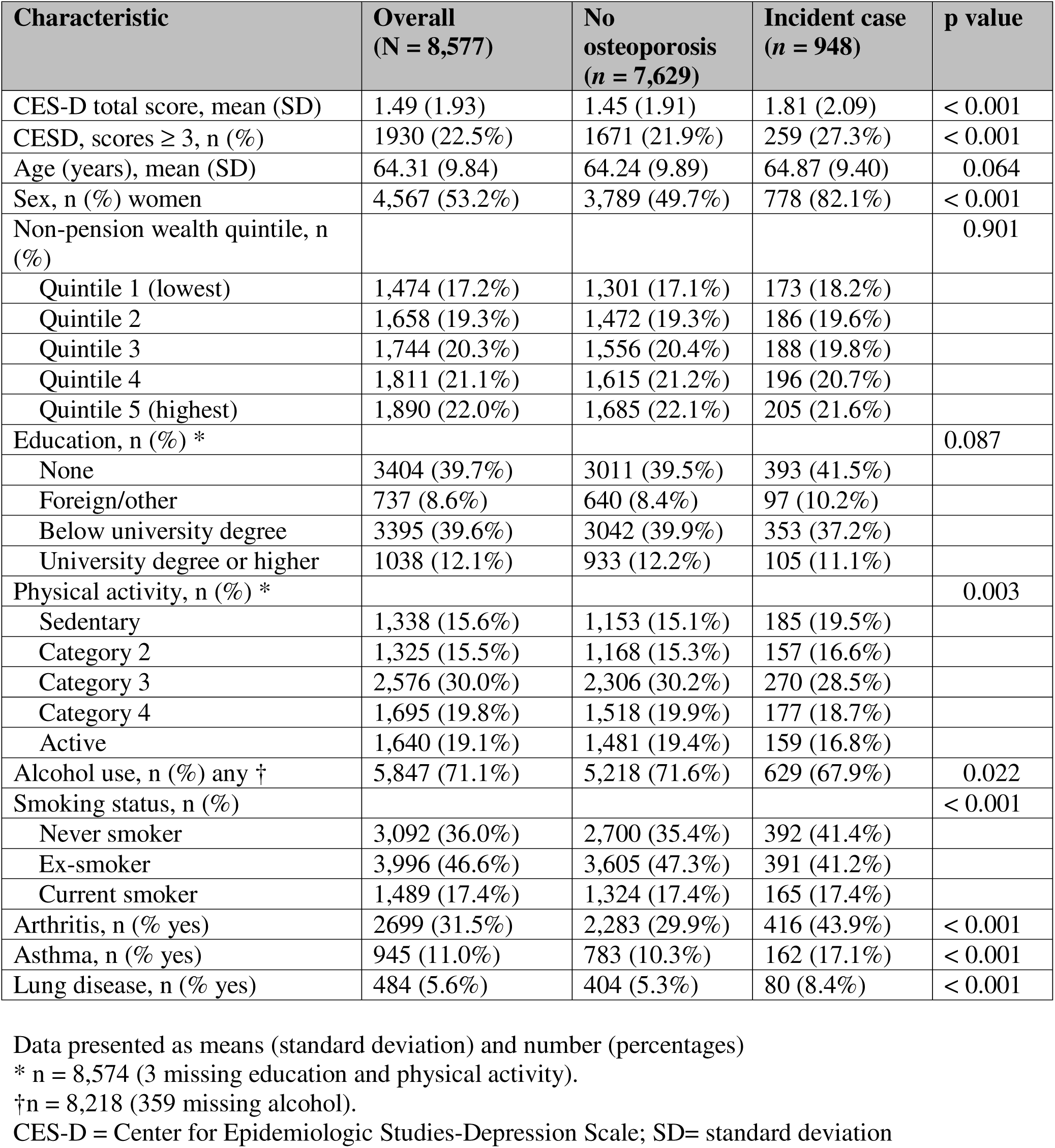
Participant characteristics (2002–03) according to incident osteoporosis status (2004–24)

| Characteristic | Overall<br>(N = 8,577) | No<br>osteoporosis<br>(n = 7,629) | Incident case<br>(n = 948) | p value |
| --- | --- | --- | --- | --- |
| CES-D total score, mean (SD) | 1.49 (1.93) | 1.45 (1.91) | 1.81 (2.09) | < 0.001 |
| CESD, scores ≥ 3, n (%) | 1930 (22.5%) | 1671 (21.9%) | 259 (27.3%) | < 0.001 |
| Age (years), mean (SD) | 64.31 (9.84) | 64.24 (9.89) | 64.87 (9.40) | 0.064 |
| Sex, n (%) women | 4,567 (53.2%) | 3,789 (49.7%) | 778 (82.1%) | < 0.001 |
| Non-pension wealth quintile, n (%) |  |  |  | 0.901 |
| Quintile 1 (lowest) | 1,474 (17.2%) | 1,301 (17.1%) | 173 (18.2%) |  |
| Quintile 2 | 1,658 (19.3%) | 1,472 (19.3%) | 186 (19.6%) |  |
| Quintile 3 | 1,744 (20.3%) | 1,556 (20.4%) | 188 (19.8%) |  |
| Quintile 4 | 1,811 (21.1%) | 1,615 (21.2%) | 196 (20.7%) |  |
| Quintile 5 (highest) | 1,890 (22.0%) | 1,685 (22.1%) | 205 (21.6%) |  |
| Education, n (%) * |  |  |  | 0.087 |
| None | 3404 (39.7%) | 3011 (39.5%) | 393 (41.5%) |  |
| Foreign/other | 737 (8.6%) | 640 (8.4%) | 97 (10.2%) |  |
| Below university degree | 3395 (39.6%) | 3042 (39.9%) | 353 (37.2%) |  |
| University degree or higher | 1038 (12.1%) | 933 (12.2%) | 105 (11.1%) |  |
| Physical activity, n (%) * |  |  |  | 0.003 |
| Sedentary | 1,338 (15.6%) | 1,153 (15.1%) | 185 (19.5%) |  |
| Category 2 | 1,325 (15.5%) | 1,168 (15.3%) | 157 (16.6%) |  |
| Category 3 | 2,576 (30.0%) | 2,306 (30.2%) | 270 (28.5%) |  |
| Category 4 | 1,695 (19.8%) | 1,518 (19.9%) | 177 (18.7%) |  |
| Active | 1,640 (19.1%) | 1,481 (19.4%) | 159 (16.8%) |  |
| Alcohol use, n (%) any † | 5,847 (71.1%) | 5,218 (71.6%) | 629 (67.9%) | 0.022 |
| Smoking status, n (%) |  |  |  | < 0.001 |
| Never smoker | 3,092 (36.0%) | 2,700 (35.4%) | 392 (41.4%) |  |
| Ex-smoker | 3,996 (46.6%) | 3,605 (47.3%) | 391 (41.2%) |  |
| Current smoker | 1,489 (17.4%) | 1,324 (17.4%) | 165 (17.4%) |  |
| Arthritis, n (%) yes | 2699 (31.5%) | 2,283 (29.9%) | 416 (43.9%) | < 0.001 |
| Asthma, n (%) yes | 945 (11.0%) | 783 (10.3%) | 162 (17.1%) | < 0.001 |
| Lung disease, n (%) yes | 484 (5.6%) | 404 (5.3%) | 80 (8.4%) | < 0.001 |

Data presented as means (standard deviation) and number (percentages)
\* n = 8,574 (3 missing education and physical activity).
†n = 8,218 (359 missing alcohol).
CES-D = Center for Epidemiologic Studies-Depression Scale; SD= standard deviation

A total of 8,577 participants were included, of whom 948 (11.1%) developed incident osteoporosis over 20 years of follow-up (mean follow-up 11.4 years, 95,554 person years). Those who developed osteoporosis had higher baseline depressive symptoms (1.81 ± 2.09 vs. 1.45 ± 1.91; *p* < 0.001) than those who did not. Incident osteoporosis was more common in women than men (9.1% vs. 2%; *p* < 0.001). Those who did and did not report osteoporosis over the study period did not differ significantly in mean baseline age (*p* = 0.064), wealth quintile distribution (*p* = 0.901) or education (*p* =0.087). The groups differed in baseline physical activity (*p* = 0.003), with those who developed osteoporosis more likely to be sedentary (19.5% vs 15.1%). Further, those who developed osteoporosis were less likely to report consuming any alcohol (67.9% vs. 71.6%; *p* = 0.022) and were more likely to be a never smoker (41.4% vs. 34.7%; *p* < 0.001). Those who reported osteoporosis over the study period were significantly more likely to have arthritis, asthma and lung disease at baseline (all *p* < 0.001).

### 3.2 Depressive symptoms and incident osteoporosis

The Kaplan-Meier curve (Figure 1) showed lower osteoporosis-free survival in participants with raised depressive symptoms at baseline (CES-D ≥ 3; unadjusted HR 1.52, 95% CI 1.32–1.75, p < 0.001). Over 95,554 person-years of follow-up, 948 incident cases of osteoporosis occurred, an overall incidence of 9.92 per 1,000 person-years. Incidence was higher in participants with raised depressive symptoms at baseline (13.52 per 1,000 person-years) than in those without (9.02 per 1,000 person-years). After adjustment for age, sex and wealth, raised depressive symptoms were associated with a higher hazard of incident osteoporosis (HR 1.24, 95% CI 1.07–1.44, *p* = 0.004).

**Figure 1:**
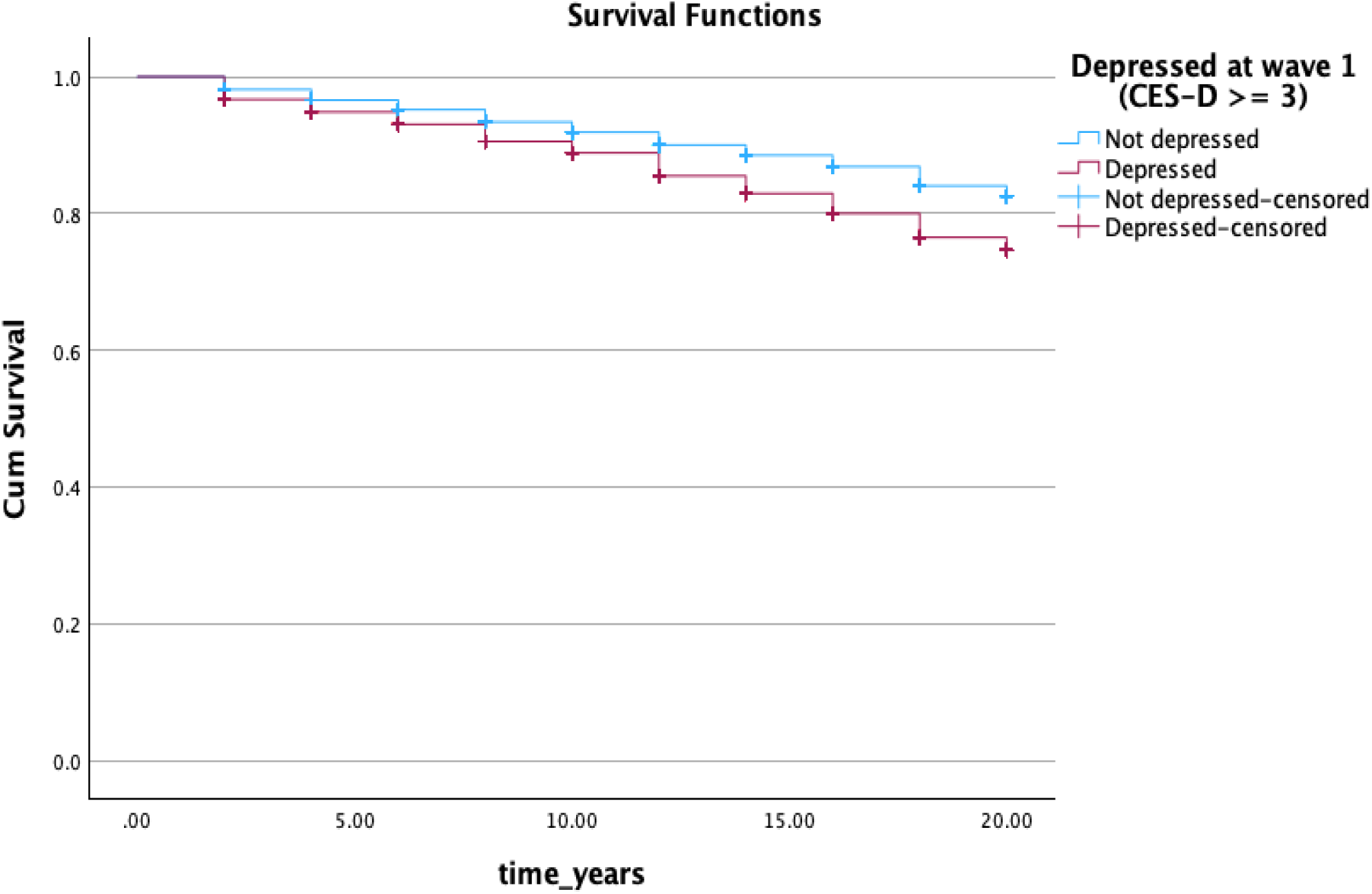
Kaplan-Meier survival curves for time to osteoporosis by baseline depressive symptom status in the English Longitudinal Study of Ageing (2002-2024) CES-D = Center for Epidemiologic Studies-Depression Scale

Each one-point increase in baseline CES-D score was associated with a higher hazard of incident osteoporosis (Table 2) adjusted for age, sex and wealth (HR 1.07, 95% CI 1.04–1.11, *p*<0.001). Age was a significant predictor of osteoporosis, with greater incidence in older participants (HR1.03; 95% CI 1.03–1.04, *p*<0.001). Female participants were significantly more likely to develop osteoporosis (HR 3.91; 95% CI 3.31–4.62 *p*<0.001). As a sensitivity check, models were re-run excluding osteoporosis cases at wave 2 (*n=* 189) to address potential reverse causation. Restricting to participants with follow-up beyond 2 years (N = 7,368; 759 events) the hazard ratio was unchanged (HR 1.07, 95% CI 1.03–1.11) *p* < 0.001). The proportional hazards assumption was not violated for this model (*p* = 0.165).

**Table 2:** Cox proportional hazards regression of baseline depressive symptoms (2002-03) on incident osteoporosis (2004-24) in the overall and age-stratified samples.

| Sample | n | Events (%) | HR per CES-D point | 95% CI | p value |
| --- | --- | --- | --- | --- | --- |
| Overall sample | 8,577 | 948 (11.05%) | 1.07 | 1.04–1.11 | < 0.001 |
| 50–64 years | 4,678 | 486 (10.4%) | 1.10 | 1.06–1.15 | < 0.001 |
| 65–74 years | 2,405 | 287 (11.9%) | 1.01 | 0.95–1.08 | 0.680 |
| 75 years and older | 1,494 | 175 (11.7%) | 1.06 | 0.99–1.14 | 0.113 |

All models adjusted for sex and non-pension wealth
CES-D = Center for Epidemiologic Studies-Depression Scale; CI = Confidence Interval; HR = Hazard Ratio.

### 3.3 Effect modification by age and sex

The CES-D × sex interaction was not significant (HR 0.97, 95% CI 0.89–1.06, *p* = 0.504). However, the CES-D × age interaction was significant (HR 0.997, 95% CI 0.994–0.999, *p* = 0.045), indicating that the depressive symptoms–osteoporosis association attenuates with increasing baseline age. In age-stratified Cox models adjusted for sex and wealth (Table 2, Figure 2), the association was significant among participants aged 50–64 years (HR 1.10, 95% CI 1.06–1.14, *p* < 0.001; 486 events in 4,678 participants), but null among those aged 65–74 years (HR 1.01, 95% CI 0.95–1.08, *p* = 0.680; 287 events in 2,405 participants) and 75 years and older (HR 1.06, 95% CI 0.99–1.14, *p* = 0.113; 175 events in 1,494 participants).

**Figure 2:**
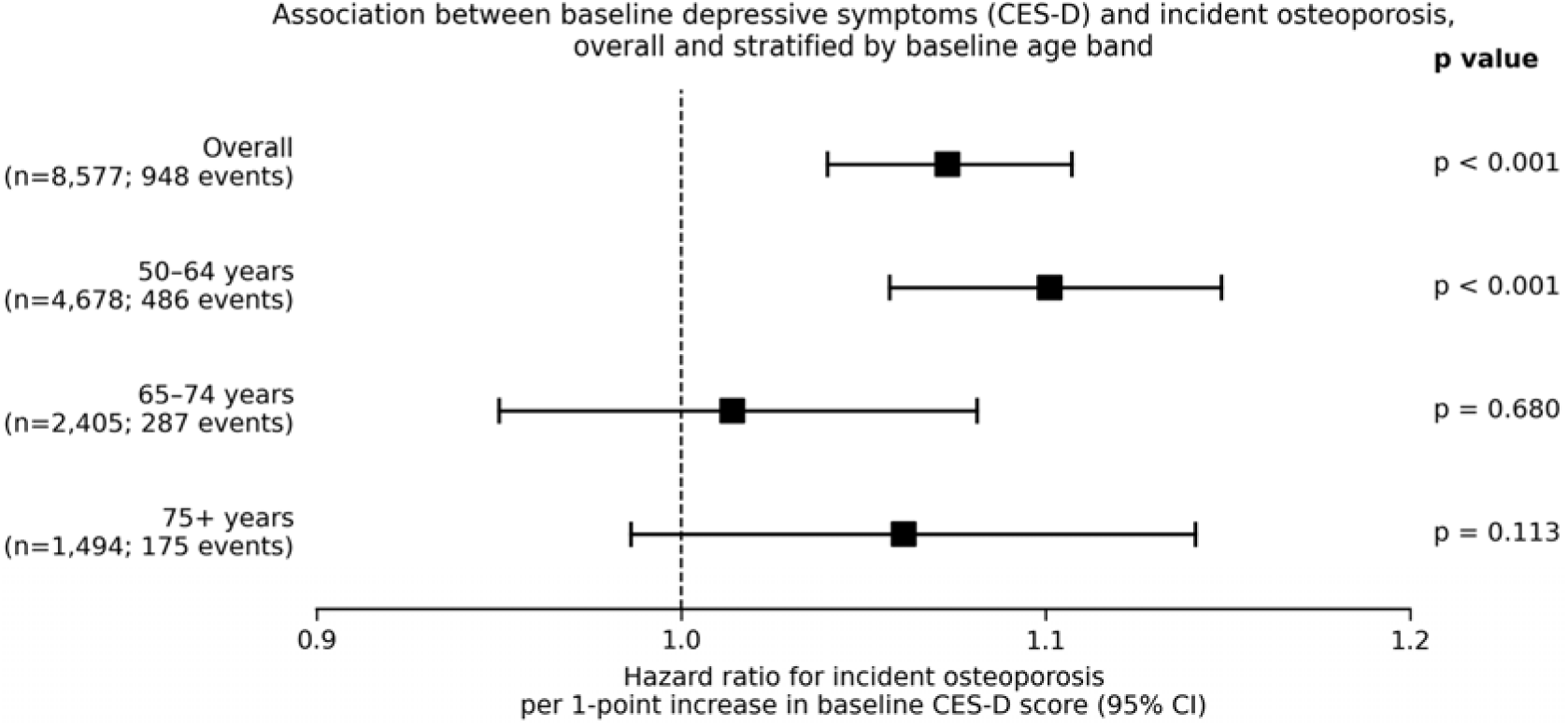
Forest plot of overall and age-stratified hazard ratios for incident osteoporosis per one-point increase in baseline CES-D score.

### 3.4 Missing data sensitivity analyses

In sensitivity analysis using multiple imputation (*n*=9,074; 20 imputations), the association between baseline depressive symptoms and incident osteoporosis was unchanged (pooled HR 1.08, 95% CI 1.05-1.11, *p* < 0.001). Imputed age-stratified results were also consistent with the main analysis, with the association significant only in those aged 50-64 years (Table 3).

**Table 3:** Overall and age-stratified Cox proportional hazards regression of baseline depressive symptoms on incident osteoporosis (multiple imputation)

| Sample | n | Events (%) | HR per CES-D point | 95% CI | p value |
| --- | --- | --- | --- | --- | --- |
| Overall sample | 9074 | 992 (10.9%) | 1.08 | 1.05-1.11 | < 0.001 |
| 50–64 years | 4,991 | 512 (10.3%) | 1.11 | 1.07–1.16 | < 0.001 |
| 65–74 years | 2,502 | 296 (11.8%) | 1.02 | 0.95–1.08 | 0.641 |
| 75 years and older | 1,581 | 184 (11.6%) | 1.06 | 0.99–1.14 | 0.094 |

Missing covariate data imputed by chained equations (20 imputations).
All models adjusted for sex and non-pension wealth
CES-D = Center for Epidemiologic Studies-Depression Scale; CI = Confidence Interval; HR = Hazard Ratio.

### 3.5 DAG informed covariate adjustment

In sequential covariate adjustment informed by a DAG (Supplementary Figure 1; *n* = 8,212, 926 incident cases), the association between baseline depressive symptoms and incident osteoporosis remained significant at every stage (Table 4). Adjusted for age, sex and education (Model 1), each one-point increase in CES-D score was associated with a higher hazard of osteoporosis (HR 1.08, 95% CI 1.05-1.12, *p* < 0.001). Additional adjustment for wealth and inflammatory conditions (Model 2) attenuated the association (HR 1.05, 95% CI 1.02-1.09, *p* = 0.003), with arthritis, asthma and chronic lung disease each independently associated with incident osteoporosis. Further adjustment for health behaviours (Model 3) left the estimate largely unchanged (HR 1.04, 95% CI 1.01-1.08, *p* = 0.010). Most of the attenuation occurred when the inflammatory conditions entered the model rather than the behaviours (a reduction in the log hazard of 38% from Model 1 to Model 2, and a further 10% from Model 2 to Model 3).

**Table 4:** Sequential covariate adjustment of the Cox proportional hazards regression between baseline depressive symptoms (2002–03) and incident osteoporosis (2004–24)

| Variable | Model 1<br>HR (95% CI) | Model 2<br>HR (95% CI) | Model 3<br>HR (95% CI) |
| --- | --- | --- | --- |
| CES-D score (per 1-point increase) | 1.08 (1.05–1.12) *** | 1.05 (1.02–1.09) ** | 1.04 (1.01–1.08) * |
| Age (per 1-year increase) | 1.03 (1.03–1.04) *** | 1.03 (1.02–1.04) *** | 1.03 (1.02–1.04) *** |
| Sex (ref: men) | 4.00 (3.37–4.74) *** | 3.83 (3.23–4.55) *** | 3.88 (3.26–4.62) *** |
| Education (per 1-category increase) | 1.00 (0.94–1.07) | 1.03 (0.97–1.10) | 1.04 (0.97–1.11) |
| Wealth (per 1-quintile increase) | - | 0.97 (0.92–1.02) | 0.98 (0.93–1.03) |
| Arthritis (ref: no) | - | 1.46 (1.27–1.67) *** | 1.45 (1.26–1.66) *** |
| Asthma (ref: no) | - | 1.43 (1.20–1.70) *** | 1.42 (1.19–1.69) *** |
| Chronic lung disease (ref: no) | - | 1.53 (1.20–1.94) *** | 1.49 (1.18–1.90) ** |
| Physical activity (per 1-category increase) | - | - | 0.94 (0.90–1.00) * |
| Alcohol use (ref: none) | - | - | 1.09 (0.94–1.26) |
| Smoking (per 1-category increase) | - | - | 1.03 (0.94–1.14) |

All models were estimated on a complete-case sample (n = 8,212; 926 incident cases).
Covariate selection was informed by a directed acyclic graph (Supplementary Figure 1).
Model 1 adjusted for age, sex and education
Model 2 additionally adjusted for wealth and inflammatory conditions (arthritis, asthma and chronic lung disease)
Model 3 additionally adjusted for physical activity, alcohol use and smoking
CES-D = Center for Epidemiologic Studies-Depression Scale; CI = Confidence Interval; HR = Hazard Ratio; Ref= Reference
\* p < 0.05; \*\* p < 0.01; \*\*\* p < 0.001.

## 4. Discussion

To our knowledge, this is the largest prospective study to examine the association between depressive symptoms and incident self-reported osteoporosis. In a representative cohort of adults living in England, higher baseline depressive symptoms were associated with an increased risk of incident osteoporosis over 20-years follow-up, independent of age, sex and wealth. Our sample size of 8,577 participants enabled subgroup analyses, where we observed significant moderation by age. Stratified analyses indicated that the association between depressive symptoms and later osteoporosis was concentrated in those aged 50-64 at baseline with no evidence of associations in those aged 65-74 or 75 and older. We did not observe any effect modification by sex.

Most prospective work linking depressive symptoms to skeletal health to date has focused on bone mineral density or incident fracture rather than self-reported osteoporosis[8, 19]. A 2018 meta-analysis prospectively associated depressive symptoms with lower bone mineral density and increased fracture risk[19]. Recent work has also prospectively associated depressive symptoms with later fracture[20]. Direct prospective evidence on incident osteoporosis is more limited, but our findings are consistent with this previous work[4, 27, 28]. A prior population-based retrospective administrative cohort study in Taiwan reported increased risk of incident osteoporosis among adults with clinical depression diagnoses, with the strongest associations in mid-life[27]. A 2026 ELSA-based trajectory analysis in a subset of 2,816 participants selected for complete four-wave depression data reported associations between depressive symptom patterns and later osteoporosis[28]. Our findings extend this emerging longitudinal evidence by examining baseline depressive symptoms in a larger representative cohort with up to 20 years of follow-up, and by characterising age modification of the association. Our results are consistent with prior work in ELSA showing depressive symptoms predict a range of chronic health conditions including osteoporosis at mid- and older age over 10 year follow-up[4].

Research in other representative cohorts has associated depressive symptoms with prevalent osteoporosis[14–16, 18]. However, cross-sectional analyses cannot determine whether depressive symptoms predict osteoporosis onset or whether depressive symptoms reflect a psychological response to osteoporosis diagnosis. Therefore, our prospective findings add to the literature in suggesting that depressive symptoms are associated with later osteoporosis onset, independent of age, sex and wealth. As our study was observational in nature, causality cannot be inferred. To address the possible risk of reverse causality, we conducted a sensitivity analysis removing cases of osteoporosis occurring within two years of baseline. The association remained consistent with the main analysis (HR 1.07 in both), suggesting that our findings are unlikely to be explained by reverse causation. Moreover, as osteoporosis is often asymptomatic until advanced, undiagnosed disease at baseline is unlikely to have generated depressive symptoms. The effect was also constant over time (suggested by the proportional hazards assumption being upheld), which we would not expect if reverse causation were driving the association. The association between depressive symptoms and incident osteoporosis was concentrated in adults aged 50–64 years at baseline, with null associations at older ages. Several pathways may contribute to this age-specific pattern. The 50–64 age band encompasses the perimenopausal and early postmenopausal window in women, a period of accelerated bone loss[29] during which depressive symptom-related biological stressors, including hypothalamic-pituitary-adrenal (HPA) axis activation, inflammation, as well as adverse health behaviours[8], may compound this loss. Comparable bone loss occurs in mid-life men through gradual testosterone decline[8], which may explain why our associations were modified by age but not by sex. Mid-life vulnerability to depressive symptom-related effects on bone may therefore reflect a shared mechanism in women and men rather than one driven exclusively by the menopausal transition. However, information on menopausal status was not collected at wave 1 of ELSA, so we were unable to test this pathway. Differential detection may also contribute; greater healthcare contact around the menopause, including breast screening from age 50 may increase the likelihood that osteoporosis is identified in this age band.

Several explanations may account for the null association in those aged 65 and over. First, older adults with depressive symptoms are at higher risk of dying from cardiovascular disease[30] and other conditions, potentially before osteoporosis becomes evident, weakening the observed association. However, we did not have access to linked ELSA mortality data to test this competing risk. Second, in ELSA those who are retained in the sample over time are healthier than those who drop out[26], so older adults who remained disease-free despite depressive symptoms may represent a selectively resilient subgroup. Third, the older subgroups were smaller with fewer events, reducing power to detect associations.

The mechanisms through which depressive symptoms may increase osteoporosis risk remain to be elucidated, but our findings point more towards biological than behavioural pathways. In our analyses, smoking, physical inactivity, and alcohol consumption together attenuated the depressive symptoms-osteoporosis association by around 10%, suggesting that these behaviours only account for a small part of the link. We did not have baseline data on BMI which is associated with both higher depressive symptoms[31] and, through greater mechanical loading, higher bone mineral density[32], so it may confound the depressive symptom to osteoporosis association in either direction. We cannot exclude residual confounding by adiposity, and the association we report here should not be interpreted as fully robust to this. Research is needed to establish whether the depressive symptom-osteoporosis relationship presented here is independent of body composition. Baseline diet, and other unmeasured behavioural factors may have contributed further attenuation we were unable to account for. Future work could explore the depression polygenic score available in a subsample of ELSA (participants from 2013-2014 onwards) as a genetic instrument[33, 34], triangulating observational estimates against an approach based on different causal assumptions.

Another possibility is that direct biological pathways link depressive symptoms and osteoporosis. Inflammation is involved in the development of osteoporosis [12]. Depressive symptoms are also associated with chronic low-grade inflammation including raised interleukin-6 and C-reactive protein[13], which may accelerate bone resorption through pro-inflammatory cytokine effects on osteoclast activity[12]. The pattern we observed as covariates were added is consistent with such a pathway as the largest reduction in the association occurred when inflammatory conditions (arthritis, asthma and chronic lung disease) entered the model rather than when health behaviours were added, and each condition was independently associated with incident osteoporosis. Because these conditions share inflammatory mechanisms with depressive symptoms and are themselves linked to bone loss[35, 36], adjusting for them may remove part of the pathway of interest rather than only confounding, so this step is better read as over-adjustment consistent with a shared inflammatory pathway than as evidence that comorbidity explains the association. However, as these conditions were measured concurrently with depressive symptoms, we cannot formally separate mediation from shared common causes. Further, depressive symptoms are known to disrupt the HPA axis, resulting in dysregulated cortisol output, which may drive bone loss by inhibiting bone formation and promoting bone resorption[8]. Sustained activation of these stress-related biological systems through chronic depressive symptoms may contribute to dysregulation of bone homeostasis. Antidepressant medication, for which we lacked data, may also contribute, as some antidepressants affect bone metabolism and have been linked to reduced bone density[37].

This study has several strengths, including the use of a nationally representative sample, a validated measure of depressive symptoms[23] and a 20-year follow-up period. We add to a sparse literature using self-reported clinical diagnoses of osteoporosis rather than bone mineral density or fracture, capturing a related but distinct aspect of bone health reflecting health-service interaction for bone health. Our large sample size enabled subgroup analyses, suggesting age modification of the depressive symptoms-osteoporosis association.

Our study also had limitations. Our data were observational, so we cannot infer causality. We did not have access to linked ELSA mortality data, so it was not possible to account for death as a competing risk. Our brief depressive symptom measure does not reflect the complexity of depressive symptoms or clinical depression diagnoses. Further, we only assessed depressive symptoms at wave 1, meaning this measure could reflect transient rather than chronic/episodic depressive symptoms. Our osteoporosis outcome was based on self-report rather than objective records, which may underestimate the number of true cases, as only those who have interacted with healthcare are captured. This may have also introduced surveillance bias as individuals with depressive symptoms consult primary care more frequently, they may be more likely to be referred for assessment and thus diagnosed, potentially inflating the observed association. Osteoporosis status was ascertained biennially, so the precise timing of onset between waves was unknown. Treating interval-censored event times as exact may introduce bias in survival estimates, although this is likely to be modest for conditions such as osteoporosis that develop slowly over time.

Missing data are common in population cohorts[22] and our primary analysis excluded participants with missing data. As the excluded group were generally more disadvantaged on characteristics associated with both depressive symptoms and osteoporosis (i.e., less wealthy and less healthy), complete-case estimates may be conservative relative to the wider population. However, sensitivity analyses using multiple imputation produced results consistent with the primary analysis, suggesting our findings were not materially impacted by missing data. We did not have data on antidepressant medication, which has been related to osteoporosis[37]. We were unable to include health-related factors such as BMI and diet as these were not assessed at wave 1. We did not account for ethnicity in our analyses due to high missingness at wave 1. Data from later waves[4, 22] indicate there are few ethnic minority participants in ELSA, which limits the generalisability of our findings to more diverse populations.

### 4.1 Conclusions

This study highlights depressive symptoms as a potential risk factor for osteoporosis over 20 years of follow-up. Associations were concentrated in those aged 50-64, offering the possibility that mid-life screening for depressive symptoms or psychosocial interventions may be relevant for bone health. It is unclear whether policies or interventions to address depressive symptoms in mid-life could help prevent the onset of osteoporosis. Further work is required to understand the potential causal nature of the depressive symptom-osteoporosis relationship, as well as underlying mechanisms, in particular biological pathways considering the lack of attenuation by health behaviours reported here.

## Ethical approval

The English Longitudinal Study of Ageing received ethical approval from the London Multicentre Research and Ethics Committee (MREC/01/02/91), in accordance with the World Medical Association Declaration of Helsinki. All participants provided informed consent at the time of data collection. As a secondary analysis of an existing dataset, no additional consent was required for this study.

## Data availability

Data from the English Longitudinal Study of Ageing are freely available to download from the UK Data Service (https://ukdataservice.ac.uk/).

## Author contributions

RAH formulated the research question, analysed and interpreted the data, and wrote the manuscript. JBG, MR, LP, AR and SN interpreted the data and reviewed and revised the manuscript. All authors approved the final version of the manuscript. RAH is the guarantor of this work.

## References

1. Willers C, Norton N, Harvey NC, et al (2022) Osteoporosis in Europe: a compendium of country-specific reports. Arch Osteoporos 17:23. 10.1007/s11657-021-00969-8

2. Al-Sari UA, Tobias J, Clark E (2016) Health-related quality of life in older people with osteoporotic vertebral fractures: a systematic review and meta-analysis. Osteoporos Int 27:2891–2900. 10.1007/s00198-016-3648-x

3. Deparment of Health and Social Care (2022) Women’s Health Strategy for England. In: GOV.UK. https://www.gov.uk/government/publications/womens-health-strategy-for-england/womens-health-strategy-for-england. Accessed 19 Jan 2026

4. Poole L, Steptoe A (2018) Depressive symptoms predict incident chronic disease burden 10 years later: Findings from the English Longitudinal Study of Ageing (ELSA). Journal of Psychosomatic Research 113:30–36. 10.1016/j.jpsychores.2018.07.009

5. Kivimäki M, Steptoe A (2018) Effects of stress on the development and progression of cardiovascular disease. Nat Rev Cardiol 15:215–229. 10.1038/nrcardio.2017.189

6. Hackett RA, Steptoe A (2017) Type 2 diabetes mellitus and psychological stress - a modifiable risk factor. Nat Rev Endocrinol 13:547–560. 10.1038/nrendo.2017.64

7. Ng CYH, Tay SH, McIntyre RS, et al (2022) Elucidating a bidirectional association between rheumatoid arthritis and depression: A systematic review and meta-analysis. Journal of Affective Disorders 311:407–415. 10.1016/j.jad.2022.05.108

8. Mezuk B, Eaton WW, Golden SH (2008) Depression and osteoporosis: epidemiology and potential mediating pathways. Osteoporos Int 19:1–12. 10.1007/s00198-007-0449-2

9. Moayyeri A (2008) The Association Between Physical Activity and Osteoporotic Fractures: A Review of the Evidence and Implications for Future Research. Annals of Epidemiology 18:827–835. 10.1016/j.annepidem.2008.08.007

10. Yoon V, Maalouf NM, Sakhaee K (2012) The effects of smoking on bone metabolism. Osteoporos Int 23:2081–2092. 10.1007/s00198-012-1940-y

11. Maurel DB, Boisseau N, Benhamou CL, Jaffre C (2012) Alcohol and bone: review of dose effects and mechanisms. Osteoporos Int 23:1–16. 10.1007/s00198-011-1787-7

12. Ginaldi L, Di Benedetto MC, De Martinis M (2005) Osteoporosis, inflammation and ageing. Immun Ageing 2:14. 10.1186/1742-4933-2-14

13. Raison CL, Capuron L, Miller AH (2006) Cytokines sing the blues: inflammation and the pathogenesis of depression. Trends Immunol 27:24–31. 10.1016/j.it.2005.11.006

14. Deng S, Xie R, Zhao W, et al (2026) Cross-national evidence on frailty-depression multi-trajectories and osteoporosis risk: Findings from the ELSA and HRS cohorts. Maturitas 204:108790. 10.1016/j.maturitas.2025.108790

15. Ma M, Liu X, Jia G, et al (2022) The association between depression and bone metabolism: a US nationally representative cross-sectional study. Arch Osteoporos 17:113. 10.1007/s11657-022-01154-1

16. Chen K, Wang T, Tong X, et al (2024) Osteoporosis is associated with depression among older adults: a nationwide population-based study in the USA from 2005 to 2020. Public Health 226:27–31. 10.1016/j.puhe.2023.10.022

17. Zhang W, Tang Y, Chen L, et al (2025) Depressive Symptoms and Osteoporosis in Middle-Aged and Older Adults: A Cross-Sectional Analysis of NHANES and HRS Data. Geriatr Orthop Surg Rehabil 16:21514593251357525. 10.1177/21514593251357525

18. Godde K, Courtney MG, Roberts J (2024) Psychological Disorders Linked to Osteoporosis Diagnoses in a Population-Based Cohort Study of Middle and Older Age United States Adults. Gerontologist 64:gnae027. 10.1093/geront/gnae027

19. Wu Q, Liu B, Tonmoy S (2018) Depression and risk of fracture and bone loss: an updated meta-analysis of prospective studies. Osteoporos Int 29:1303–1312. 10.1007/s00198-018-4420-1

20. Zhao P, Ying Z, Yuan C, et al (2024) Shared genetic architecture highlights the bidirectional association between major depressive disorder and fracture risk. Gen Psych 37:. 10.1136/gpsych-2023-101418

21. Sialino LD, van Oostrom SH, Wijnhoven HAH, et al (2021) Sex differences in mental health among older adults: investigating time trends and possible risk groups with regard to age, educational level and ethnicity. Aging & Mental Health 25:2355–2364. 10.1080/13607863.2020.1847248

22. Steptoe A, Breeze E, Banks J, Nazroo J (2013) Cohort profile: the English Longitudinal Study of Ageing. Int J Epidemiol 42:1640–1648. 10.1093/ije/dys168

23. Radloff LS (1977) The CES-D Scale A Self-Report Depression Scale for Research in the General Population. Applied Psychological Measurement 1:385–401. 10.1177/014662167700100306

24. He J, Deng S, Xie G, et al (2026) The association between body mass index and osteoporosis, with consideration of sex differences: a systematic review and dose-response meta-analysis. BMC Musculoskelet Disord 27:362. 10.1186/s12891-026-09675-3

25. White IR, Royston P (2009) Imputing missing covariate values for the Cox model. Stat Med 28:1982–1998. 10.1002/sim.3618

26. Banks J, Muriel A, Smith JP (2011) Attrition and health in ageing studies: Evidence from ELSA and HRS. Longitudinal and life course studies 2:. 10.14301/llcs.v2i2.115

27. Lee CW-S, Liao C-H, Lin C-L, et al (2015) Increased risk of osteoporosis in patients with depression: a population-based retrospective cohort study. Mayo Clin Proc 90:63–70. 10.1016/j.mayocp.2014.11.009

28. Liao C-C, Liao K-R, Li J-M (2026) Distinct Impacts of Somatic and Cognitive-Affective Depressive Symptom Trajectories on Osteoporosis Risk in Adults Aged 50 Years and Older. Clinical Gerontologist 0:1–13. 10.1080/07317115.2026.2675036

29. Greendale GA, Sowers M, Han W, et al (2012) Bone mineral density loss in relation to the final menstrual period in a multiethnic cohort: results from the Study of Women’s Health Across the Nation (SWAN). J Bone Miner Res 27:111–118. 10.1002/jbmr.534

30. White J, Zaninotto P, Walters K, et al (2016) Duration of depressive symptoms and mortality risk: The English Longitudinal Study of Ageing (ELSA). The British Journal of Psychiatry 208:337–342. 10.1192/bjp.bp.114.155333

31. Luppino FS, de Wit LM, Bouvy PF, et al (2010) Overweight, obesity, and depression: a systematic review and meta-analysis of longitudinal studies. Arch Gen Psychiatry 67:220–229. 10.1001/archgenpsychiatry.2010.2

32. Lloyd JT, Alley DE, Hawkes WG, et al (2014) Body mass index is positively associated with bone mineral density in US older adults. Arch Osteoporos 9:175. 10.1007/s11657-014-0175-2

33. Ajnakina O, Cadar D, Steptoe A (2020) Interplay between Socioeconomic Markers and Polygenic Predisposition on Timing of Dementia Diagnosis. J Am Geriatr Soc 68:1529–1536. 10.1111/jgs.16406

34. Khatun T, Norton S, Hackett RA (2026) The interplay between polygenic susceptibility and discrimination on depressive symptoms: a prospective cohort study. Aging & Mental Health 30:481–491. 10.1080/13607863.2025.2569654

35. Dam T-T, Harrison S, Fink HA, et al (2010) Bone mineral density and fractures in older men with chronic obstructive pulmonary disease or asthma. Osteoporos Int 21:1341–1349. 10.1007/s00198-009-1076-x

36. Rauner M, Bozec A (2026) Mechanisms of osteoclast activation in inflammatory bone loss in rheumatoid arthritis. Nat Rev Rheumatol 22:42–61. 10.1038/s41584-025-01323-9

37. Rajha HE, Abdelaal R, Charfi K, et al (2025) Examining depression, antidepressants use, and class and their potential associations with osteoporosis and fractures in adult women: Results from ten NHANES cohorts. Journal of Affective Disorders 369:1223–1232. 10.1016/j.jad.2024.10.114

