## Supplementary Figures 1 and 2 for "Depressive symptoms and incident osteoporosis: Findings from a 20-year follow up of the English Longitudinal Study of Ageing"

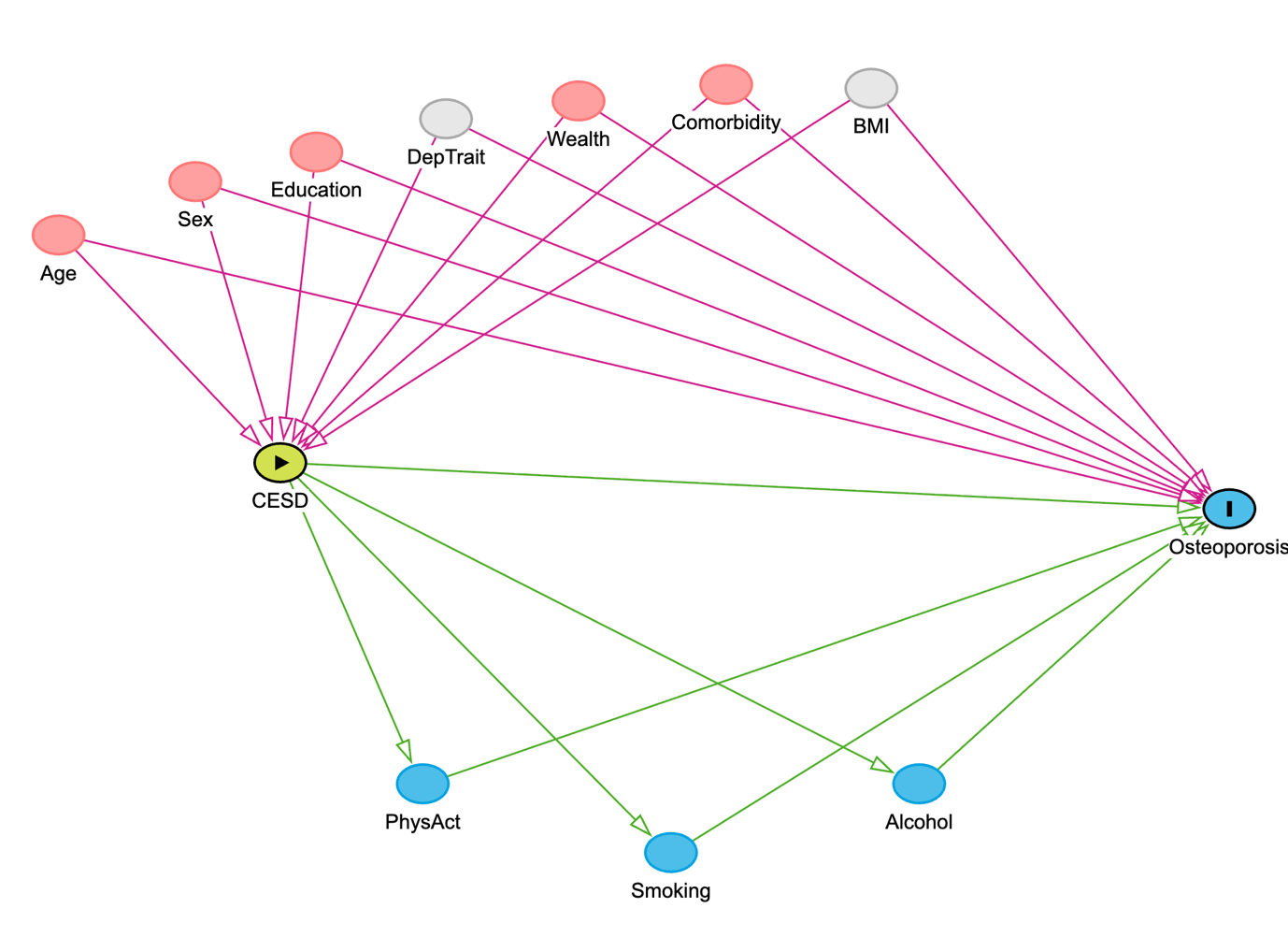
**Supplementary Figure 1.** Directed Acyclic Graph (DAG) of the hypothesised casual pathway between depressive symptoms and incident osteoporosis

BMI: Body Mass Index; CESD: Centre for Epidemiologic Studies Depression Scale; Comorbidity: Inflammatory conditions (arthritis; chronic lung disease; asthma); DepTrait: history/trait depressive symptoms; PhysAct: physical activity.

The green node denotes the exposure, depressive symptoms (CESD). Pink nodes indicate measured baseline confounders (age, sex, education, wealth, and inflammatory comorbidity). Grey nodes indicate unmeasured confounders (history/trait depressive symptoms and BMI). Blue nodes denote behavioural pathways (physical activity, smoking and alcohol consumption) Arrows indicate the assumed direction of causal influence between variables.

**Supplementary Figure 2. Flow diagram of participants included in and excluded from the analyses**

| ELSA wave 1 (2002–03)  (*n* = 12,099) |  |  |
| --- | --- | --- |
| **↓** | **⟶** | Excluded:  Aged under 50 years  (*n* = 577) |
| Aged ≥ 50 years at baseline  (*n* = 11,522) |  |  |
| **↓** | **⟶** | Excluded:  Prevalent osteoporosis at wave 1  (*n* = 565) |
| Free of osteoporosis at wave 1  (*n* = 10,957) |  |  |
| **↓** | **⟶** | Excluded:  Missing baseline CES-D data  (*n* = 426) |
| With complete baseline CES-D data  (*n* = 10,531) |  |  |
| **↓** | **⟶** | Excluded:  Missing wealth covariate data  (*n* = 281) |
| Baseline eligible sample  (*n* = 10,250) |  |  |
| **↓** | **⟶** | Excluded:  Did not provide follow-up data  (*n* = 1,673) |
| Analytic sample for survival analysis  (*n* = 8,577)  (948 incident cases over follow-up) |  |  |

CES-D = Center for Epidemiologic Studies-Depression Scale; English Longitudinal Study of Ageing
